# Neuroinflammation in the amygdala is associated with recent depressive symptoms

**DOI:** 10.1101/2022.12.19.22283678

**Authors:** Wei Zhang, Jerrel Rutlin, Sarah A. Eisenstein, Yong Wang, Deanna Barch, Tamara Hershey, Ryan Bogdan, Janine Bijsterbosch

## Abstract

**Background:** Converging evidence suggests that elevated inflammation may contribute to depression. Yet, the link between peripheral and neuro-inflammation in depression is unclear. Here using data from the UK Biobank (n=11,512), we estimated associations among depression, C-reactive protein as a measure of peripheral inflammation (CRP), and neuroinflammation as indexed by diffusion-basis spectral imaging-based restricted fraction (DBSI-RF).

**Methods:** DBSI-RF was derived from diffusion-weighted imaging data for whole-brain gray matter (global-RF), and regions of interest in bilateral amygdala (amygdala-RF) and hippocampus (hippocampus-RF), and CRP was estimated from blood (serum) samples. Self-reported recent depression symptoms were measured using a 4-item assessment. Linear regressions were used to estimate associations between CRP and DBSI-RFs with depression, while adjusting for the following covariates: Age, sex, body mass index, smoking, drinking, and medical conditions.

**Results:** Elevated CRP was associated with higher depression symptoms (r=0.03, p<0.001) and reduced global-RF (r=-0.03, p<0.005). Higher amygdala-RF was associated with elevated depression – an effect resilient to added covariates and CRP (t=2.53, β=0.02, p<0.05). Interestingly, this association was stronger in individuals with a lifetime history of depression (t=3.02, β=0.07, p<0.005) than in those without (t=2.32, β=0.03, p<0.05). Associations between global-RF or hippocampus-RF with depression were not significant, and no DBSI-RF indices indirectly linked CRP with depression (i.e., mediation effect).

**Conclusion:** Peripheral inflammation and DBSI-RF neuroinflammation in the amygdala are independently associated with depression, consistent with animal studies suggesting distinct pathways of peripheral and neuro-inflammation in the pathophysiology of depression, and with investigations highlighting the role of the amygdala in stress-induced inflammation and depression.

## INTRODUCTION

Major depressive disorder (MDD) is a worldwide health issue with profound emotional, cognitive, and economic consequences (1,2). Building upon meta-analytic studies that elevated inflammation is associated with depression (3,4), convergent evidence supports the theory that inflammation is a potential etiological factor contributing to the development of depression (5– 14). For example, immunological challenges (e.g., flu vaccine in humans, lipopolysaccharide administration in non-human animals; interferon treatment in both) induce depressive symptoms (15,16), whereas anti-inflammatory treatment reduces depressive symptoms (17–19). Furthermore, inflammation is prospectively associated with future depression, even when considering depression at baseline (4,13) and baseline depression has been prospectively associated with future elevations in inflammation (20), suggestive of a bidirectional relationship that may contribute to the widespread chronicity and recurrence among individuals suffering from depression (3). Although a multitude of mechanisms may contribute to the relationship between peripheral inflammation and depression, the putative depressogenic effects of inflammation are often attributed to neuroinflammation (21). However, there are limited studies of neuroinflammation in humans, and the available evidence is mixed due to small sample sizes in positron emission tomography (PET) studies on depression (22–24). While studies of microglial activity using translocator protein (TSPO) will be informative (25), increasing evidence suggests that biological correlates of complex behavioral phenotypes like depression are characterized by small effects (26), highlighting the need to extend neuroinflammatory research into larger datasets. Here, we leverage the large-scale UK Biobank dataset (UKB; analytic N=11,501) to examine the relationship between neuroinflammation index and depressive symptoms.

Although neuroinflammation cannot be directly measured in humans in vivo, currently available neuroimaging methods including PET, single photon emission computed tomography (SPECT), and magnetic resonance imaging (MRI) have been utilized for indirect assessment. Recent evidence from PET and functional MRI techniques indicates that putative neuroinflammation in depression might be most relevant for subcortical regions that subserve reward, motivation, and emotion processes. For example, increased plasma CRP concentration was associated with decreased functional connectivity between the amygdala and ventromedial prefrontal cortex in patients with depression comorbid with anxiety disorder relative to healthy controls (27), and depressive symptom severity was correlated with significant increases in TSPO total density in the hippocampus in patients compared to healthy controls (28). Interestingly, alterations in structure and function of the hippocampus and amygdala are also the most prominent neuroimaging markers for major depression disorder (29–32). These two subcortical regions are involved in a wide range of cognitive and emotional processes including emotion regulation, memory and stress, and impairment in these functions are often evident in individuals with depression (33–35). It remains understudied, however, whether inflammation in these regions may contribute to depressive symptom severity, and whether disruptions in these regions may potentially mediate the impact of peripheral inflammatory response on depression symptomatology. In this study, we used diffusion basis spectrum imaging (DBSI), a novel non-invasive technique, to estimate putative neuroinflammation in these regions of interest (ROI) from diffusion-weighted imaging (DWI) data.

The DBSI approach is an extension of standard diffusion tensor imaging (DIT) that models the entire DWI signal as a linear combination of multiple anisotropic diffusion tensors and a spectrum of isotropic diffusion components (36,37). In comparison to DTI that describes water movement parallel and perpendicular to axon tracts, the DBSI approach has the advantage to improve specificity and sensitivity of diffusion properties by resolving multiple-tensor water diffusion resulting from axon injury, demyelination and inflammation (36). In particular, DBSI-based restricted fraction (DBSI-RF), the metric of interest in this study, provides a putative indicator of neuroinflammation-related cellularity (i.e., the state of having cells), where higher values indicate increased immune cell infiltration relative to lower values (36). DBSI-RF has shown robust correlations with cell nuclei counts in brain tissues in a mouse model of autoimmune encephalomyelitis, with stain-quantitated cellularity from post-mortem human CNS tissues and has been found to be greater in individuals with inflammation-related diseases such as multiple sclerosis, obesity, HIV and Alzheimer’s disease than in controls (38–42). These studies suggest that DBSI-RF can be employed as a proxy measure for inflammation-related cellularity in the brain.

For this study, our goal was to assess putative inflammation-related cellularity (i.e., DBSI-RF) specifically in the hippocampus, amygdala, and whole-brain gray matter in relation to depression. We further explored whether putative neuroinflammation mediates the association between peripheral inflammation and depression.

## METHODS

### Participants

The UK Biobank (UKB) is a large-scale study (N>500,000 participants) designed to examine the genetic, environmental, biological, and behavioral correlates of broad-spectrum health outcomes and related phenotypes (43). For the present study we considered data from 16,182 participants who completed the baseline session (i.e., the first assessment center visit between 2006-2010), initial neuroimaging acquisition (i.e., the third assessment center visit from 2014-2019) and a web-based assessment session (2016-2017) of the UKB study.

Data were excluded for the following reasons: 1) mismatch between self-reported and genetic sex (N=130), 2) missing data on a variable included in this study (N=1,834), 3) marked elevation of C-reactive protein (CRP) levels (i.e., above 10gm/L; N=398) that may reflect acute infection, injury, or disease (44), and 4) diseases associated with systemic inflammation (e.g., known inflammatory disease, autoimmune disease, HIV, Hepatitis B or C; N=2,319). Thus, our final analytic sample consisted of 11,501 participants (*Table 1*). This study was conducted under the UK Biobank Application ID 47267.

**Table 1.** Demographics of study sample

|  |  | FULL | Lifetime MDD |  |  |
| --- | --- | --- | --- | --- | --- |
|  |  |  | YES | NO | All |
| **Size** | Female | 6,097 (53%) | 1,138 (67%) | 2,368 (45%) | 3,506 (50%) |
|  | Total | 11,501 | 1,693 | 5,267 | 6,960 |
| **Age** |  | 62.41 (7.41) | 61.17 (7.15) | 63.05 (7.43) | 62.59 (7.41) |
| **Race (White %)** |  | 98.37% | 99.17% | 97.80% | 98.13% |
| **Body Mass Index (kg/m<sup>2</sup>)** |  | 24.81 (2.74) | 24.75 (2.81) | 24.82 (2.70) | 24.80 (2.73) |
| **Serum C-reactive Protein (mg/L)** |  | 1.42 (1.46) | 1.43 (1.45) | 1.40 (1.44) | 1.41 (1.44) |
| **RDS** |  | 5.15 (1.68) | 5.68<sup>****</sup> (2.02) | 4.73<sup>****</sup> (1.33) | 4.97 (1.58) |
| **DBSI-RF** | Gray Matter | 0.071<br>(0.011) | 0.071<br>(0.011) | 0.071<br>(0.011) | 0.071 (0.011) |
|  | Hippocampus | 0.037<br>(0.005) | 0.037<br>(0.005) | 0.037<br>(0.005) | 0.037 (0.005) |
|  | Amygdala | 0.028<br>(0.006) | 0.028<br>(0.005) | 0.028<br>(0.006) | 0.028 (0.006) |

Mean (S.D.) are reported for each variable except for sample size and race ratio. \*\*\* indicates significant differences ( $p < 0.00001$ ) in RDS between the phenotype and the full sample such that individuals with lifetime MDD had a higher and individuals without lifetime MDD a lower RDS score than that of the full study sample.
RDS: recent depressive symptoms; DBSI-RF: diffusion basis spectrum imaging-based restricted fraction

In a subset of sample for our secondary analysis on lifetime major depressive disorder (MDD) phenotype, data of participants who met lifetime MDD criterion were excluded if they had reported other mood disorders (e.g., bipolar, schizophrenia) or on treatment or medication for antipsychotics, whereas data of participants who did not meet lifetime MDD criterion were excluded if they had a history of depression or any mental problems, or on antidepressants (see detailed exclusion criterion in *Table S2*). The resulting sample had 6,960 participants, including 1,138 participants with lifetime MDD (*Table 1*).

### Depression

The primary measure of depression used in our study was the total score of recent depressive symptoms (RDS), which was completed on the same day of the neuroimaging session. This score was a sum of 4 items (scored on a 1-4 scale where 1 = not at all, 4 = nearly every day) assessing the presence of the following self-reported depressive symptoms over the last 2 weeks: 1) depressed mood, 2) unenthusiasm/disinterest, 3) tenseness/restlessness, and 4) tiredness/lethargy. The resulting sum score ranged between 4 and 16, where higher scores indicate more frequent and severe depressive symptoms. RDS has been previously validated against several commonly used depression scales including PHQ-9 (45). Variable IDs of these items in the UKB Data Showcase are summarized in *Table S1*.

In secondary analyses, we focused on a subset of participants of lifetime MDD phenotype (46). Lifetime MDD is defined based on the short form of the Composite International Diagnostic Interview (CIDI-SF) (47) that was administered as part of the online mental health questionnaire (46). Briefly, participants were asked to identify their experiences regarding two core depression symptoms: a) depressive mood and b) loss of interest, for a period of two or more weeks. If “Yes” was replied to any of these questions, participants continued to indicate the lifetime number of depressive symptoms, and their experiences on the other DSM-IV MDD symptoms during the worst episode: (c) feelings of worthlessness, (d) tiredness, (e) difficulty concentrating, (f) suicidal thoughts, (g) changes in sleeping pattern, and (h) changes in weight. This provides a 0-8 sum score of depression symptoms over 8 symptom questions, where higher scores indicate greater endorsement of depression symptoms. Participants were also asked to indicate the frequency and duration of the core symptoms, as well as their psychological impairment (i.e., whether they interfere with their roles, life, and activities). An individual participant is considered to have lifetime MDD when their sum score ≥ 5, and experienced symptoms “almost every day” or “every day”, with a duration of “most of the day” or “all day”, and symptoms impaired psychosocial functioning “somewhat” or “a lot” (46). Following previous studies (48,49), we further applied exclusion criterion to ensure phenotype specificity to depression (e.g., excluding participants reporting bipolar disorder or schizophrenia). Details are summarized in Supplemental Materials (*Table S2*). As shown in *Table 1*, the mean depression symptom scores in individuals with lifetime MDD were significantly higher, whereas that in individuals without lifetime MDD were lower, compared to the full study sample. Of note, a small subset of participants (N=2,044) completed the online mental health questionnaire up to 1.8 years after the neuroimaging session.

### Inflammation Indices

Peripheral inflammation was indexed by baseline serum CRP level (mg/L), which was measured by an immunoturbidimetric high-sensitivity analysis on a Beckman Coulter AU5800 (see Biomarker assay quality procedures for more details: https://biobank.ctsu.ox.ac.uk/showcase/showcase/docs/biomarker_issues.pdf). The blood sample used for CRP assessment was collected at the baseline visit, which was on average 7.6 years prior to the acquisition of DWI data and depression assessment in this study sample. We also generated Indices of neuroinflammation using DWI data (see details below in *DBSI and Neuroinflammation Index*).

### Imaging acquisition and processing

This study made use of DWI and T1-weighted structural MRI data (i.e., FreeSurfer-based segmentation) that were processed and generated by an image-processing pipeline developed and ran on behalf of UK Biobank (50). Specifically, DWI data (2×2×2 mm^3^) were acquired using a multi-shell approach with two b-values (b = 1000 and 2000 s/mm^2^). For each diffusion-weighted shell 50 diffusion-encoding directions were acquired. Preprocessing of DWI data included eddy currents and head motion corrections, outlier slice correction and gradient distortion correction (50).

High-resolution T1 structural MRI data (1×1×1 mm^3^) was acquired with an in-plane acceleration sequence. The preprocessed T1 images (i.e., after removing the skull and correcting for gradient distortion) were further processed with FreeSurfer, and the outputs (e.g., images, surface files and summary outputs) are made available for download from the UKB Data Showcase. Details of the acquisition protocols, image processing pipeline, and derived imaging measures can be found in the UK□Biobank□Imaging□Documentation (https://www.fmrib.ox.ac.uk/ukbiobank/) and the study by Miller et al. (51).

### DBSI and Neuroinflammation Index

To index inflammation in the brain (neuroinflammation), we derived restricted fraction (RF) from diffusion-weighted imaging data, using DBSI (36,39,41).

We focused on the hippocampus and amygdala, our ROIs, and obtained subject-specific segmentation images of these two subcortical regions from the UKB FreeSurfer outputs, which were generated by FreeSurfer’s aseg tool (52). We then extracted mean DBSI-RF from these ROIs, separately for the left and right hemispheres, using the FreeSurfer segments as masks and FSL function “fslmeants” (53). The extracted RF values were then averaged within each ROI as they showed moderate-to-high left-right correlations (r’s>0.68). Additionally, we derived a global index of neuroinflammation in whole-brain gray matter (i.e., global-RF), using the FSL FAST-based gray matter segmentation (54) as the mask. The resulting RFs of whole-brain gray matter, hippocampus and amygdala were included in separate statistical analyses.

### Covariates

Age, sex, body mass index (BMI), ethnicity, smoking and drinking statuses were included as covariates to adjust for potential confounds. Additionally, we included a composite measure to index medical conditions that combined information about cancer diseases, non-cancer diseases, medical operations, and medications. Covariate measurements and their variable IDs in the UKB database are described in *Supplemental Table S1*.

### Statistical Analysis

To investigate whether DBSI-RF relates to depression symptoms, we conducted a series of linear regression analyses. Whenever significant associations were observed between DBSI-RF, depression, and CRP, we conducted planned mediation analyses to estimate whether DBSI-RF may indirectly link CRP to depression, using PROCESS macro modeling tool that is implemented in R (i.e., R package *bruceR* (55)). These analyses were performed separately for whole-brain gray matter, hippocampus, and amygdala, with adjustment for covariates (i.e., adjusted for both dependent and mediator variables). In case of significant effects of neuroinflammation indices, we further tested in the follow-up regression analyses to determine whether their effects were independent of CRP concentration. In mediation analyses, CRP concentration and RDS were included as independent and dependent variables, respectively. We conducted these analyses for the full study sample, as well as for the selected subset of participants based on lifetime MDD definition. The regression models for lifetime MDD participants further included an interaction term of lifetime MDD status (“Yes” vs. “No”) with DBSI-RFs. To ensure robust statistical results, we conducted diagnostic analyses to test multi-collinearity of independent variables in each regression model that returned significant effects of interest. Specifically, we calculated a generalized variance inflation factor (GVIF) score for each variable under investigation and investigated whether any observed effects might be biased due to high GVIF values (i.e., high collinearity).

FDR corrections were applied to all regression analyses to account for multiple testing.

## Results

### Study Sample Demographics

Participants were predominately White, middle-old aged (mean age=62.41), and about half were female. On average, participants scored 5.15 (SD=1.68) on RDS questions, indicating relatively low levels of depression. Lifetime MDD participants showed similar demographic characteristics as the full study sample. Detailed demographic information for the study samples is summarized in *Table 1*.

### Associations Between Neuro-inflammation, Peripheral Inflammation, and Depression

#### Peripheral Inflammation and Depression

Consistent with a recent study leveraging a larger UK Biobank sample unconstrained by the need for neuroimaging data (56), CRP was positively associated with recent depressive symptoms in our study sample (t=3.45, β=0.04, p_fdr_<0.005). This association remained following covariate inclusion (t=2.57, β=0.02, p_fdr_<0.05).

#### Associations with Neuroinflammation Estimates

DBSI-RF metrics in whole-brain gray matter, hippocampus and amygdala were associated with each other (β=0.09-0.41, p_fdr_’s<0.005), with the strongest correlation between amygdala-RF and hippocampus-RF. Interestingly, only global-RF was correlated with CRP concentration (t=-3.64, β=-0.03, p_fdr_<0.001); this negative association remained significant with adjustment for all covariates (β=-0.02, p_fdr_<0.05). We did not find association of hippocampus-RF (β=-0.01, p_fdr_=0.17), or of amygdala-RF (β=-0.01, p_fdr_=0.12) with CRP.

In line with our hypotheses, RF of the amygdala was associated with significantly more depressive symptoms with the adjustment for covariates (t=2.53, β=0.02, p_fdr_<0.05; *Figure 1A*), and further inclusion of CRP concentration (t=2.58, β=0.02, p_fdr_<0.05). Neither the global-RF (t=1.87, β=0.02, p_fdr_=0.10), nor the hippocampus-RF (t=1.23, β=0.01, p_fdr_=0.21) showed such an association. Summary statistics for these associations between neuro-inflammation estimates and depressive symptoms can be found in Supplemental Materials (*Table S3*).

**Figure 1.**
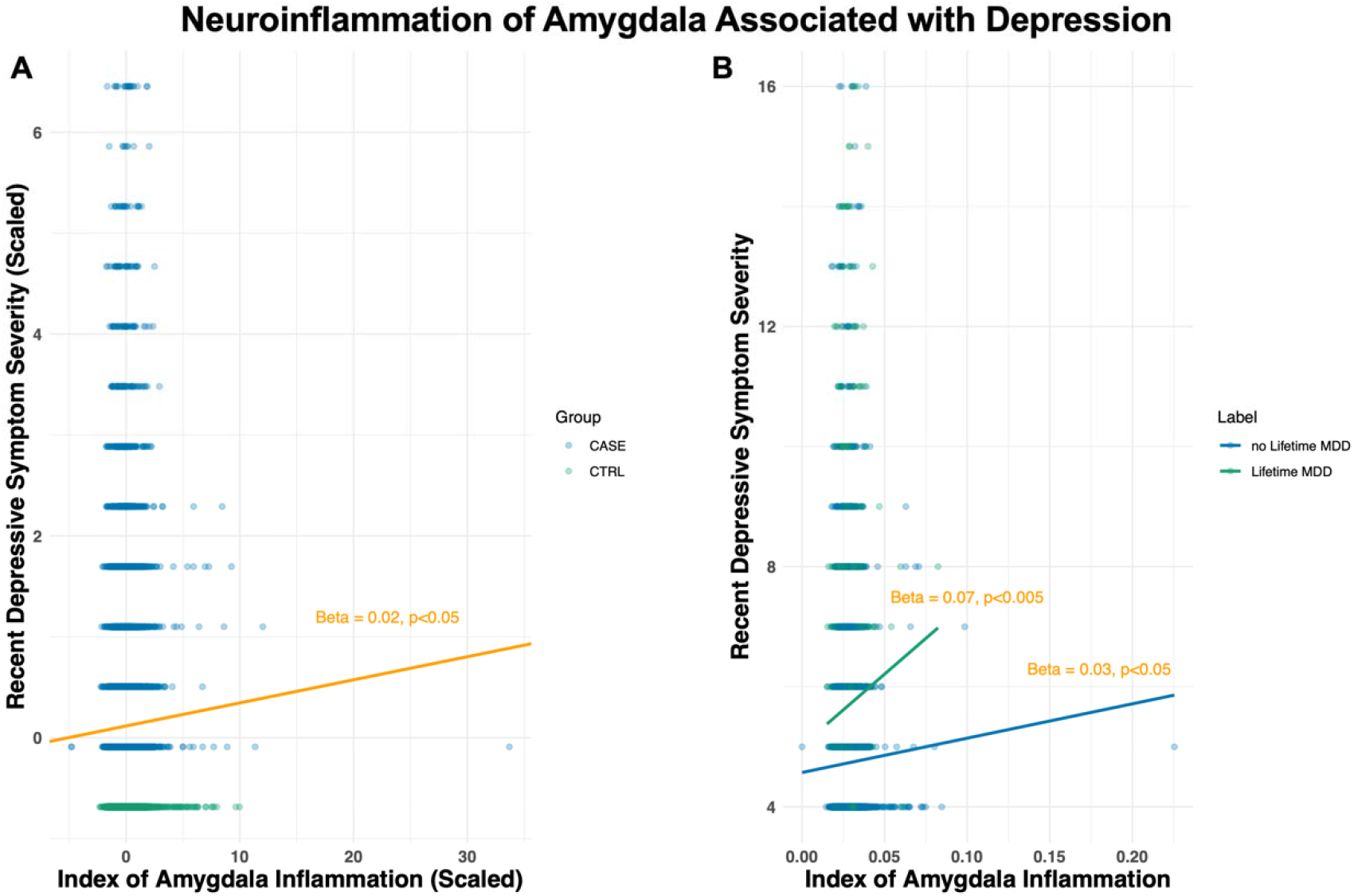
Robust association between the amygdala inflammation index and depression. Restricted fraction (RF) – the index of neuroinflammation -in the amygdala was associated with depression: it predicted recent depressive symptom severity in the full study sample (A) and showed a stronger predictive effect on participants with lifetime MDD than those without lifetime MDD (B). Note, all results remained significant with removal of the data point in the “no Lifetime MDD” group that showed abnormally large value (more than 3 S.D. above mean) of amygdala-RF (see *Robustness of Effects*).

As CRP was only significantly associated with global-RF, we conducted the hypothesized mediation model for these variables. However, we did not find evidence supporting a mediation role of global-RF on the association between peripheral inflammation and depression (indirect effect *ab*: Z=-1.33, p=0.18).

### Secondary Analyses for Lifetime MDD Phenotype

Results from our secondary tests on lifetime MDD phenotype broadly recapitulated the findings observed with recent depressive symptoms described above, with minor differences (*Table S4*). Briefly, although we did not observe differences in CRP concentration nor in global-RF or hippocampus-RF between lifetime MDD and non-lifetime MDD participants (p’s>0.3), we found a significant interactive effect of group (lifetime MDD versus no lifetime MDD) and amygdala-RF on depression symptoms (t=2.59, β=0.08, p_fdr_<0.05; *Figure 1B*). This effect was robust against all covariates and further inclusion of CRP concentration (t=2.6, β=0.08, p_fdr_<0.05). The follow-up tests show that amygdala-RF had a stronger relation to depression symptoms in individuals with (t=3.02, β=0.07, p<0.005) than in those without lifetime MDD (t=2.32, β=0.03, p<0.05). These effects remained with further adjustment for CRP effect (p’s<0.03).

### Robustness of Effects

As some of the predictors and covariates are correlated (*Figure S1*), we conducted follow-up tests to examine whether the collinearity between predictors and covariates might have biased the observed results. Our findings show that the observed associative effects of CRP, and DBSI-RF of the amygdala on depression symptoms are robust against multicollinearity, as indicated by very low generalized variance inflation factor (GIVF) values for each independent variable in the regression models (i.e., all below 1.12; see full list in *Table S5*).

Additionally, because nearly half of the participants in the full study sample reported no symptoms (N=5,592) – “Not at all” to all four symptom questions, we further conducted a logistic regression analysis to test whether the neuroinflammation indices of the amygdala could also predict group identities (i.e., control group = reporting zero symptoms). We replicated the amygdala-RF effect showing that amygdala neuroinflammation could differentiate two groups of participants based on their depressive symptoms (Z=3.06, β=0.06, p<0.005), with specificity of 0.62 and sensitivity of 0.56. This predictive effect remained when CRP concentration was considered in addition to the inclusion of all covariates (Z=3.09, β=0.06, p<0.005).

Furthermore, to ensure that effects of amygdala-RF were not driven by outliers (see *Figure 1*), we repeated the analyses for the full study sample and lifetime MDD, respectively, after excluding the outliers above 3 S.D. of the sample mean (N=91 in the full study sample; N=59 in the lifetime MDD sample). Results remained consistent with minor changes in coefficients (main effect in full study sample: t=2.96, β=0.03, p<0.005; interactive effect with lifetime MDD label: t=2.1, β=0.03, p<0.05).

Lastly, we repeated the analysis with the amygdala-RF by lifetime MDD label interaction for a subset of participants (N=6,019), whose assessment of lifetime MDD phenotype was performed prior to that of recent depressive symptom (i.e., ensuring lifetime MDD as a history in time). We observed an even stronger interactive effect on depression symptoms (t=2.78, β=0.11, p<0.001), which was driven by significant amygdala-RF impact on depression symptomatology in individuals with lifetime MDD (N=842; t=2.83, β=0.10, p<0.005). Amygdala-RF showed no association with depression in individuals without lifetime MDD (t=1.18, β=0.03, p=0.24).

## Discussion

In the current study, we applied a novel non-invasive imaging technique to derive index measures of neuroinflammation in two a priori selected subcortical regions – the hippocampus and amygdala -that have been consistently linked to depression symptomatology (29–31,33). Our study aimed to investigate whether the derived neuroinflammation indices could explain variance in depressive symptom severity and whether such neuroinflammation might mediate the impact of peripheral inflammation on depression. Our findings demonstrate an association between neuroinflammation at brain regional levels and depressive symptomatology.

In particular, our findings show that the putative neuroinflammation index of the amygdala has a small but significant association with depressive symptoms, especially in individuals with a lifetime history of MDD. This signal in the amygdala retained its association with depression even after adjusting for effects of demographics, lifestyles, and CRP concentration that also all showed significant associations with depressive symptoms. In light of recent investigations on the UKB dataset suggesting that the more strictly defined lifetime MDD phenotype (i.e., based on CIDI-SF;(47)) may have higher genomic and behavioral relevance specific major depression (57,58), our results highlight a direct link between neuroinflammation index (i.e., inflammation-related cellularity) in the amygdala and elevated depressive symptoms. Our findings are also consistent with a growing body of evidence showing that inflammation can contribute to depression via the stress-induced remodeling of amygdalar structure and function (59). Future studies may further investigate whether the depression-associated putative neuroinflammation are accompanied by other alterations in structure and functions in this subcortical region and in connected regions known to be involved in depression, and whether stress-related experiences may modulate such associations.

Contrary to our hypotheses, however, we did not observe an association between hippocampus-RF and depression symptomatology even though the DBSI-RFs of the hippocampus and amygdala were moderately associated (β=0.41) and these two subcortical regions are both considered highly relevant to depression (29–31,33). These results are broadly in line with recent studies that linked peripheral inflammation markers (e.g., CRP) to brain structure and function and suggested that specific brain circuitry might be more vulnerable to inflammation impact in individuals with depression (27,60,61). It awaits to be tested whether other brain regions such as regions from the reward circuitry (e.g., ventral striatum, caudate nucleus, and putamen) may exhibit greater sensitivity to depression-related inflammation. Alternatively, previous studies have linked peripheral inflammation markers with depression symptom clusters (62) or subtypes (63), suggesting that inflammatory underpinnings of depression may be better characterized by symptom dimensions (64). In this study, we used a sum score of four items to indicate the overall severity of recent depressive symptoms. The limited number of questions potentially restricted explorations on depression with a multidimensional approach. Future investigations may consider utilizing depression assessment instruments that allow for quantifying symptom dimensions and examining the associated inflammatory profiles.

In line with the literature (6,9), we also found a positive association between the baseline CRP concentration and depressive symptom severity. This effect remained significant after controlling for physical illness, demographic characteristics including age, sex, BMI, and smoking and drinking status. In contrast to our expectations, however, we did not find evidence suggesting that DBSI-RFs in the whole-brain gray matter, hippocampus, or the amygdala could mediate the CRP relations to depression symptomatology. In fact, the amygdala-RF and baseline CRP appeared to explain complementary variance in depression symptoms in the study samples (i.e., showing significant effects separately in one multiple linear regression model). Notably, the collection of blood samples for baseline CRP concentration took place on average 7.6 years before DWI data acquisition. This large time difference might have blurred the relationship between the inflammation measures in the peripheral and central nervous systems. Nevertheless, when we repeated the models including this time difference as an additional covariate, the relations of baseline CRP concentration and neuroinflammation in the amygdala with depression symptomatology remained significant (amygdala: t=2.61, β=0.02; CRP: t=2.61, β=0.02; p’s<0.01).

## Limitations

The major limitation of this study concerned the time difference of data acquisition: blood sample for assessing peripheral inflammation marker CRP was collected on average 7.6 years prior to the acquisition of imaging data and assessment of depression symptom. Although CRP is stable over time and prospectively associated with depression in older adults (13,65), this large time difference may have attenuated associations of both depression and neuroinflammation with CRP.

It should also be noted that the neuroinflammation measure (i.e., DBSI-RF) in our study only indirectly assesses inflammation-related cellularity. Although it has not been tested in individuals with depression, DBSI-RF has been linked to increased cell nuclei numbers and activated microglia in several neuroinflammatory conditions (36,37,40,41), and associated with Alzheimer’s disease and obesity (38,39,42), for which neuroinflammation is hypothesized to play a critical role (66,67). Our finding of the amygdala-RF in relation to depressive symptoms demonstrates a potential contribution of neuroinflammation to depression symptomatology. Lastly, our study only examined the putative neuroinflammation index in relation to depressive symptoms. It remains to be tested whether the observed amygdala-RF effect is specific to depression and not shared by other highly comorbid disorders such as anxiety.

## Conclusions

Our study applied the novel DBSI technique to derive regional putative neuroinflammation indices for whole-brain gray matter, hippocampus, and amygdala, and linked them with depression symptomatology. Although we failed to find evidence supportive of mediating effects for these neuroinflammation measures on the association between peripheral inflammation and depression, we did observe a robust and significant effect of putative neuroinflammation in the amygdala on depressive symptom severity across individuals with and without a lifetime history of MDD, but more strongly in the former group. Future studies are warranted to expand the focus of brain regions outside the hippocampus and amygdala, and to assess whether neuroinflammation in other depression-related regions (e.g., in reward circuitry) can also explain variance in depression symptomatology. With the very first evaluation of DBSI-based neuroinflammation in relation to depression symptomatology, our study provides early evidence suggesting the significance of regional neuroinflammation in depression symptom severity. These data add to a growing body of research linking inflammation with depression and demonstrating potential underlying mechanisms for depression pathogenesis.

## Data Availability

All data produced in the present work will be returned to UK Biobank upon journal publication.

## Acknowledgement

WZ is supported by McDonnell Center for Systems Neuroscience at Washington University in Saint Louis; SAE received fundings from WUSTL Diabetes Research Center Pilot & Feasibility Award (P30 DK020579 (SAE)), Neuroimaging Laboratory Research Center Innovation Matching Funds (SAE), the Mallinckrodt Institute of Radiology Pilot Award (SAE). RB received support from: R01AG061162, R21AA027827, R01 DA054750, U01DA055367. JB is supported by the NIH (NINDS R34 NS118618; NIMH R01 MH128286) and the McDonnell Center for Systems Neuroscience.

